# Accessing Prosthetic and Rehabilitation Services After Lower Limb Amputation in Cambodia: A Qualitative Study of Service Users and Families

**DOI:** 10.64898/2026.01.12.25342916

**Authors:** Maggie Donovan-Hall, Chantel Ostler, Ratana Sann, Cheryl Metcalf, Peter Worsley, Sisary Kheng, Sam Simpson, Carson Harte, Alex Dickinson

## Abstract

**Purpose:** While research has improved understanding of prosthetic use in Cambodia, little is known about how individuals with lower limb amputation and their families experience access to prosthetic and rehabilitation services. This qualitative study explores these experiences to inform equitable and contextually appropriate service provision.

**Methods:** A qualitative study was conducted using semi-structured interviews with adult lower-limb prosthetic users and their family members attending prosthetic and rehabilitation services provided by a Non-Governmental Organisation. The study design and research materials were co-produced with the Cambodian partner research team to ensure cultural appropriateness, accessibility, and acceptability. Data were analysed inductively using reflexive thematic analysis.

**Results:** Three overarching thematic categories were identified, each comprising three themes: (1) heavy reliance on a well-functioning prosthesis, (2) accessing and using prosthetic and rehabilitation services, and (3) sources of support and impacts on quality of life. Participants described prosthetic devices as essential to mobility, independence, and social participation, while also highlighting challenges related to device durability and fit, travel and financial constraints, service continuity, and the central role of family support in enabling ongoing prosthetic use.

**Conclusions:** Prosthetic and rehabilitation services are critical to supporting life after lower limb amputation in Cambodia; however, their effectiveness is shaped by the availability of durable, well-fitting devices and services that actively address structural, social, and cultural barriers to access. Service models that are responsive, culturally sensitive, and community-oriented are required to support sustained prosthetic use holistically and to inform the future development of data-driven approaches to improving service accessibility.

## Introduction

An appropriate and functional prosthesis can confer substantial benefits for individuals with lower limb amputation (LLA), including increased mobility, greater independence, enhanced social participation, and improved overall quality of life (QoL) [1–5]. It is estimated that more than 176 million people worldwide live with limb absence, many of whom require prosthetic and orthotic (P&O) services [7]. Despite this considerable need, as many as 80–90% of individuals globally lack access to appropriate prosthetic care and resources [8]. As with other forms of rehabilitation, the unmet need for P&O services is disproportionately high in resource-limited environments (RLEs), particularly in settings affected by humanitarian disasters, armed conflict, or landmine legacies [8,9]. Consequently, substantial disparities persist in P&O service provision between RLEs and higher-income countries, or less ‘fragile’ states [8,10,11].

Limited access to prosthetic devices restricts mobility for individuals with LLA and has broader consequences beyond physical functioning. These disparities in service provision have been argued to undermine multiple human rights, including access to employment, food, shelter, and full participation in society [7,12,13]. Addressing such inequalities is therefore a fundamental component of legislation and policy governing entitlement to rehabilitation programmes [14]. International frameworks, including the United Nations Convention on the Rights of Persons with Disabilities [15] and the Agenda for Sustainable Development [16], explicitly outline obligations to improve access to mobility aids and assistive technologies through targeted research, development, and service delivery initiatives. In parallel, several reviews of P&O provision in low- and middle-income countries (LMICs) [8,9,14,17], alongside targeted empirical research [11,18], have identified priority areas for future research and intervention development, particularly in relation to improving service access and ensuring the appropriateness of assistive technologies.

Together with global policies and strategies (e.g. World Health Organisation [13,19]; United Nations [16]), this evidence base has contributed to a clearer understanding of global P&O needs within LMICs. However, it is increasingly recognised that new technologies and service models will only be effective if they are grounded in a clear understanding of the social and cultural contexts in which they are implemented [11,20,21]. Understanding the experiences of key stakeholders, particularly service users, within specific social contexts is therefore essential. Such insight helps ensure that changes to P&O service provision are appropriate, acceptable, and sustainable by aligning with local environmental, cultural, and social needs [4,11].

The importance of understanding local experiences of P&O service use in LMICs has been demonstrated by a growing body of qualitative research, including studies of lower-limb prosthetic users in South Africa [22], Nepal [23], Bangladesh [4], and Cambodia [5]. These studies provide rich, contextualised insights into the social and cultural dimensions of living with lower-limb loss, as well as the barriers and facilitators to access to rehabilitation services. Such evidence has been recognised as fundamental to identifying areas for improvement in existing P&O services and for shaping future research agendas [24], helping to ensure that services are fit for purpose.

Cambodia is a particularly important context for examining access to prosthetic and rehabilitation services. The country has a prolonged history of conflict and instability, peaking during the Khmer Rouge era (1975–1979) and continuing until the early 1990s (5). This legacy has resulted in a large population of individuals living with lower limb amputation due to conflict-related injuries and landmines, alongside a more recent increase in amputations associated with road traffic accidents and illness [25]. This evolving pattern of limb loss highlights the importance of understanding both the lived experience of using a prosthesis and accessing rehabilitation services, to ensure that prosthetic provision enables individuals to maximise functional and social outcomes.

Previous research in Cambodia has explored the everyday experiences of lower limb prosthesis users, demonstrating that successful adjustment is shaped by social interactions with other prosthesis users and highlighting the importance of social and cultural factors. This work also underscores the value of access to relevant and timely information about prosthetic services in supporting adaptation and ongoing engagement with rehabilitation [26]. In addition, a retrospective analysis of three decades of routinely collected service data has provided important insights into how prosthetic devices in Cambodia are worn, maintained, repaired, and replaced, emphasising the central role of functionality, cost, and durability in ensuring that prostheses are fit for purpose within this specific context [25]. While these studies have advanced understanding of prosthetic use in Cambodia, there remains a need for in-depth qualitative exploration of how individuals and families currently experience access to prosthetic and rehabilitation services.

This study builds on existing work by exploring the lived experiences of individuals with LLA and their family members who access prosthetic services in Cambodia. Using a qualitative approach, we draw on interviews with service users and family members to examine experiences of accessing and engaging with prosthetic and rehabilitation services within this specific context. By foregrounding these perspectives, the study aims to generate contextually grounded insights to inform the development of equitable, responsive, and sustainable prosthetic and rehabilitation services across regions.

## Materials and Methods

### Study Design and Context

A qualitative study was developed from a series of scoping and ‘Patient and Public Involvement and Engagement (PPIE)’ activities. Given the stakeholder-led, exploratory focus, it was considered that an approach independent of a theoretical and epistemological framework would provide the required flexibility [27]. We therefore employed a semi-structured interview design and applied reflexive thematic analysis as an inductive analytical approach [28,29].

### Research team

Information about the research team is provided to support reflexivity and allow readers to consider how researchers’ backgrounds may have influenced the study. The first author (XX), a health psychologist and associate professor, designed the study and led the UK and Cambodian partner teams. She has over 20 years’ experience conducting qualitative research with people following limb loss across diverse social and cultural contexts. She was supported by XX, a consultant clinical academic physiotherapist with over 20 years’ experience in prosthetic rehabilitation and 10 years’ research experience, including qualitative enquiry. The Cambodian partner team comprised XX and XX, both with backgrounds in social work in Cambodia. XX is a lecturer with interests in disability and ageing and, at the time of the study, was a welfare officer at the INGO Exceed Worldwide, which provides free prosthetic and orthotic services across Southeast Asia.

### Governance and Ethics Procedures

Full ethical approval was granted by the ‘Ministry of Health, National Ethics Committee for Health Research, Cambodia’ (No 311, NECHR) and ‘Faculty of Engineering & Physical Sciences’ at the University of Southampton (ERGO45577).

### Patient and Public Involvement and Engagement (PPIE)

As part of the exploratory nature of the work, initial scoping interviews with Cambodian prosthetic service users and health care professionals were carried out by the lead author (XX). This PPIE work aimed to inform the study design and participant information and to engage key individuals from the local clinical team as research partners, who would help co-design the study and undertake local data collection. A bespoke programme was developed to train the Cambodian research partner team (XX and XX) in qualitative research. The training was delivered over five days and included interactive teaching sessions, role-play, practice opportunities, and pilot interviews. These sessions also involved co-developing semi-structured interview schedules to ensure cultural and social appropriateness. Different sets of semi-structured interview schedules were developed for the client and family participant groups. These activities not only ensured the research project was acceptable and accessible for participants in the local setting but also contributed to building research capacity within the local clinical team.

### Participants and Recruitment

A convenience sample of 12 prosthetic users with lower limb amputation and 12 family members was recruited from three Exceed Worldwide prosthetic clinics in Phnom Penh, Sihanoukville, and Kampong Chhnang, all of which provide services free of charge.

Eligibility was determined using the inclusion and exclusion criteria outlined in Table 1, and all participants were able to communicate in Khmer and understand the study aims. Informed by PPIE feedback, multiple recruitment strategies were used to enhance accessibility and accommodate differing literacy levels. Study posters were displayed in each clinic, and clinic staff identified eligible prosthetic users from patient records and informed them about the study during clinic visits or by telephone. Prosthetic users were approached first, and, with their consent, family members were subsequently invited to participate and provided with study-specific information.

**Table 1.** Participant Inclusion and exclusion criteria.

|  | **Participant Group** | **Inclusion Criteria** | **Exclusion Criteria** |
| --- | --- | --- | --- |
| (1) | Clients | - Over the age of 18<br>- Lower limb amputation<br>- Current or previous use of a lower limb prosthetic | - Other complex/life-threatening health issues (for ethical and patient burden issues)<br>- Unable to communicate in English or Khmer<br>- Unable to provide written informed consent |
| (2) | Family members | - Over the age of 18<br>- Has a family member with lower limb loss | - Family member with limb loss is not happy for them to take part in the study<br>- Unable to communicate in English or Khmer<br>- Unable to provide written informed consent |

Before providing written consent, all potential participants had the opportunity to ask the research team any questions and were given at least 24 hours to decide whether to participate in the study.

### Data Collection

XX and XX conducted interviews in settings chosen by participants, including the three prosthetic clinics and participants’ homes. All interviews were digitally audio-recorded, and contextual field notes were written to support data interpretation. Interviews with prosthetic users lasted 20-60 minutes, whereas interviews with family members ranged from 15 to 40 minutes.

Interviews were transcribed verbatim in Khmer and subsequently translated into English. In line with best practice for qualitative research translation [29,30], a subset of transcripts was back-translated into Khmer to assess translation accuracy. All identifying information, including names of centres and specific locations, was removed from transcripts, and pseudonyms were assigned to all participants to ensure anonymity.

### Data Analysis

To explore participants’ experiences, transcripts were analysed using reflexive thematic analysis within an inductive, data-driven framework, allowing for systematic identification and interpretation of patterns of meaning in the data [26–28,31]. Client and family member transcripts were initially analysed separately, with themes later reviewed and integrated due to substantial overlap in findings. To ensure that cultural and social meanings were accurately interpreted, analysis was undertaken collaboratively by researchers based in the UK (XX, XX) and research partners in Cambodia (XX). Analysis began with familiarisation through repeated reading of transcripts (XX, XX), followed by the co-production of initial codes using hand coding. Themes were developed through constant comparison and iterative discussion, supported by ongoing reference to the raw data (XX, XX, XX) [28,31]. Emerging themes were then organised and documented, with supporting codes and illustrative quotations (XX, XX). Draft themes were reviewed with Cambodian research partners to ensure cultural relevance before being shared with the broader research team for refinement and agreement [26–28,31]. Throughout the process, reflexive diaries were maintained (XX, XX) and discussed regularly within the research team to examine the assumptions and perspectives that shaped the study.

## Results

### Participants

Twenty-four participants were recruited for the study (12 clients and 12 family members). The client group comprised 11 males and one female, with ages ranging from 23 to 62 years (mean age = 48 years). The 12 family members were related to the client in various ways, including spouse and children, grandchildren (over the age of 18), and, in one case, a neighbour rather than a family member who participated. Details of each participant and their family member are shown in Table 2.

**Table 2.** Participant characteristics: age, gender, level of amputation, occupation and family members’ relationship.

|  | Client* | Age range | Gender | Level of Amputation | Cause of amputation | Family* | Relationship to client participant |
| --- | --- | --- | --- | --- | --- | --- | --- |
| (1) | Kheang | 30-39 | Male | TF | Machinery accident | Ing | Mother |
| (2) | Vong | 50-59 | Male | TF | Traffic accident | Siha | Daughter |
| (3) | Vuthy | 50-59 | Male | TF | Land mine | Heang | Relationship not stated |
| (4) | Eoun | 50-59 | Male | TT | Land mine | Soriya | Relationship not stated |
| (5) | Ty | 60-69 | Male | TT | Infection following snake bite | Domlai | Grandson |
| (6) | Thorn | 50-59 | Male | TT | Land mine | Chany | Daughter |
| (7) | Sokhea | 30-39 | Female | TF | Traffic accident | Leakna | Niece |
| (8) | Ratanak | 20-29 | Male | TF | Traffic accident | Sreysros | Wife |
| (9) | Sarik | 50-59 | Male | TT | War accident | Sarun | Neighbour |
| (10) | Souy | 40-49 | Male | TF | Fall | Sophy | Brother |
| (11) | Rany | 30-39 | Male | TF | War accident | Nareach | Wife |
| (12) | Samey | 40-49 | Male | TF | Traffic accident | Sreynai | Wife |

\*Pseudonyms used throughout
\*TT = Transtibial and TF = Transfemoral

### Themes

Themes were identified and grouped into three main categories as shown in figure 1. Each of the categories contains discrete themes which will be explored in detail below and illustrated with quotes from clients and their family members.

**Figure 1.**
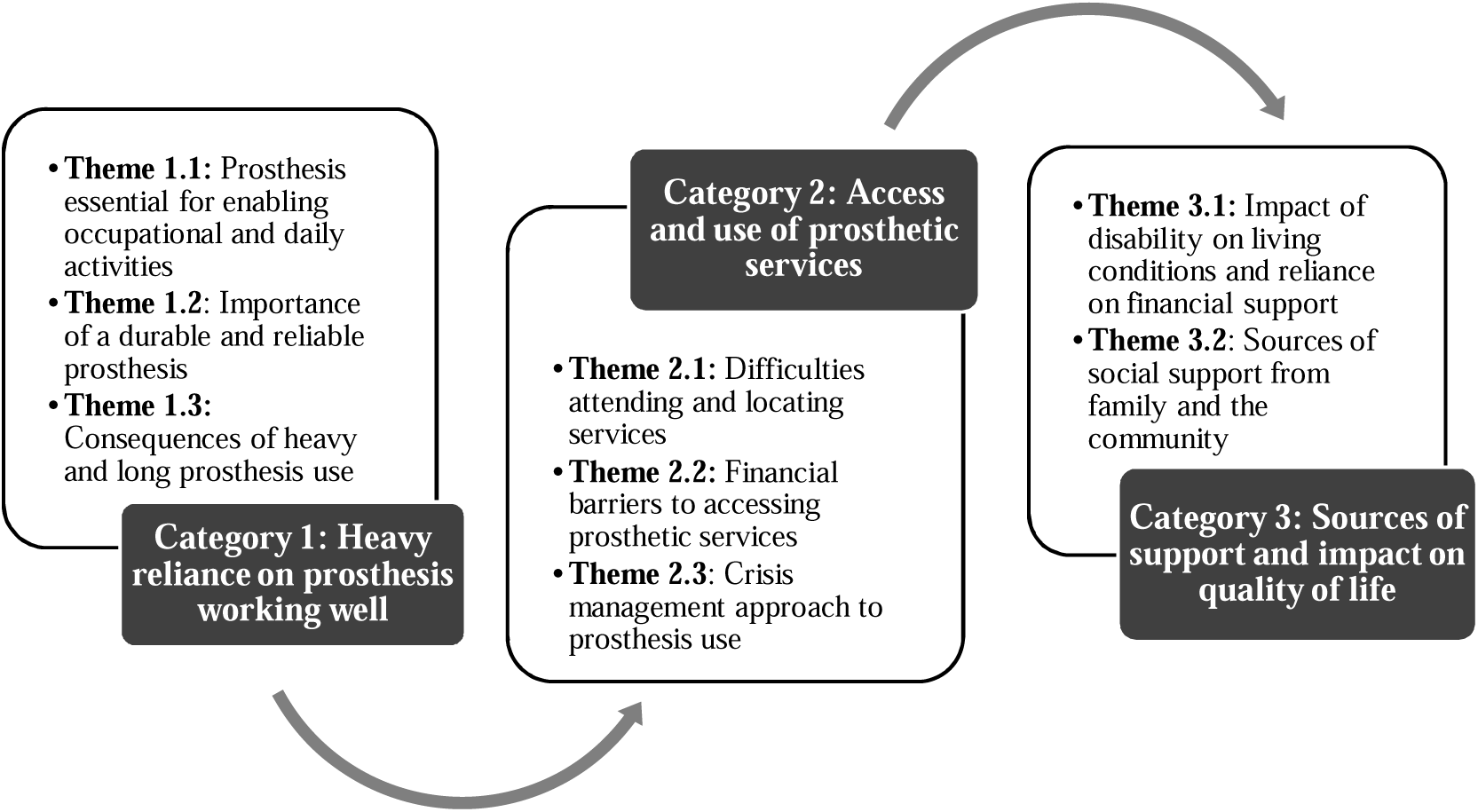
Categories and themes identified from interviews with clients and family members.

### Category 1: Heavy reliance on the prosthesis working well

Throughout the interviews both prosthetic users and their families talked about how important the prosthetic limb was to them. This heavy reliance understandably raised significant concerns and implications for people’s lives if the prosthesis failed to function adequately. Three themes explore this relationship.

#### Theme 1.1: Prosthesis is essential for enabling occupational and daily activities

Participants described the importance of the prosthesis in enabling all their daily activities, ranging from housework, personal care, essential travel for work and healthcare. In particular, their prosthesis enabled employment.

> ***‘****I really need this leg, not only for work but also for my family businesses, including taking my children to school, working and walking’ (Rany, Client, lines 86-86)*

This is also highlighted by the high use discussed by Rany and Sokhea:

> ‘*‘I wear it all the time. Though at home, I do take it off, I use a crutch but if I have to go to work, I wear it from 7am to 7pm’ (Sokhea, Client, Lines 85-86)*

> *‘I use it all the time and it has helped me. I don’t think I can live without it’ (Rany, Client, Lines 80-81)*.

It was often reported that without the prosthesis, participants would be unable to work and therefore generate income to support themselves and their families:

> ‘*I think if we do not wear any leg, we cannot work at all but if we wear it, we can go anywhere, we can earn money to support children and the wife and the family. (Soy, Client, Lines 58-59)*.

In addition to enabling day-to-day activities and supporting paid employment, it was clear that the prosthesis was also essential for enabling participation in valuable family roles. This was considered important by both clients and their family members, as this participation contributed to the wider family dynamics and functioning. This is described by Domlai referring to his Grandfather:

> *‘Most of the housework is done by him because me and my aunty always away from home. Aunty is busy from morning until evening…. I also have no free time because of my pre-class study, therefore all of the housework is done by him..’ (Domlai, Client, lines 21 – 24)*.

Some participants discussed the experience of not being able to wear their prosthesis and having to use crutches instead. Crutch use made the completion of daily activities more challenging. Ty described how crutches restrict what he could do as both hands are not free:

> ‘*That all kind of works are required me to wear the leg. It is needed, otherwise need to use crutches. Walking with crutches is very difficult to do all type of housework because the hands are not free, need to hold crutches’ (Ty, Client, Lines 154-156)*.

Some participants perceived that using crutches would highlight their disability to employers who would think that they were incapable of more physical roles or needed to prove how they could manage using crutches first:

> *‘[using crutches]… but only the light work or if they used to see me doing it they will let me do it. But if they never see they will not let me do it’ (Kheang, Client, Lines, 259-262)*.

This reliance on crutches when not being able to wear a prosthesis was also described by a relative and appeared to be associated with a higher risk of falling:

> *‘..whenever he goes around he need to use crutches on one side as you see. As walked with one leg and crutches is not easy sometime, he can slip and falls when he gets up…’ (Ing, Client, Lines, 63-65)*.

The prosthesis’s utility in enabling activity, participation, and income, combined with the challenges described here of using crutches, appeared to lead participants to use their prosthesis for many hours at a time.

> *‘I wear it all the time, except during the night. I wear it from waking up in the morning, until I go to bed at night’ (Samey, Client, Lines 66-67)*.

These descriptions of reliance on the limb for participation and heavy prosthetic use were discussed alongside the need for reliable and durable limbs. This is explored in the next theme.

#### Theme 1.2: The importance of a durable and reliable prosthesis

Participants appeared to value the durability and reliability of their prosthetic limbs and this was expressed in comments they made about the length of time their prosthesis would last, with participants often wearing the same limb for many years:

> *‘If it’s good, I can use it up to 10 years each. But sometimes, our stumps shrinks, or we gained weight, so it can be tight or loose, it will hurt and we need to replace it’ (Samey, Client, Lines 60-61)*.

Participants described the durability of their prosthesis in various ways, such as the expected average lifespan, the number of prostheses they had used since their amputation, or the duration of use of their current prosthesis. Examples of these descriptions are shown in Table 3.

**Table 3.** Participant descriptions of prosthetic durability.

| Participant | Level of amputation | Description of prosthetic durability |
| --- | --- | --- |
| Kheang | Transfemoral | *'last approximately 2 years'* |
| Vong | Transfemoral | *'about every 2 years'* |
| Vuthy | Transfemoral | *'6 legs in 26 years'* |
| Eoun | Transtibial | *'4 legs in 18 years'* |
| Ty | Transtibial | *'3 legs in 10 years, but had same leg for five years'* |
| Thorn | Transtibial | *'3 prosthesis in 20 years'* |
| Sokhea | Transfemoral (primary user) | *'first leg lasted 8 months before requiring a repair'* |
| Ratanak | Transfemoral | *'current leg lasted approximately one year'* |
| Sarik | Transtibial | *'1 leg last 3 to 5 years'* |
| Souy | Transfemoral | *'same leg for 4 years'* |
| Rany | Transfemoral | *'6 legs in 20 years'* |
| Samey | Transfemoral | *'3 legs in 13 years'* |

As highlighted by Sarik, the length of time a prosthesis lasted was put into context by the amount of use on a daily basis:

> *‘We can use 3 to 4 year if we take care of it and use it since 6am to 10pm only taking off when I’m resting’ (Sarik, Client, Lines 30 – 31)*.

Durability and reliability appeared to be particularly important because of the challenging environmental conditions and physical roles that the participants in this study were employed in, such as construction work and farming:

> *‘…..during that time I was the farmer working on the farm the whole day from the morning until evening, need to stand in the muddy condition which was not easy to lift the leg out from the mud as the sound side. And I needed to stand a lot working on that rice grain.’ (Ty, Client, Lines 123-137)*.

The impact of heavy prosthetic use and challenging environmental conditions are described in the next theme.

#### Theme 1.3: Consequences of heavy and long prosthetic use

Prosthesis use in Cambodia, as described by the participants in this study, appeared to be characterised by heavy reliance on the prosthesis, resulting in long hours of use in often challenging environmental conditions. The combination of these factors seemed to cause problems for Cambodian limb wearers, such as sweating, swelling and pain. It was frequently described as leading to limb fitting issues, potential skin breakdown and inability to wear the limb:

> *‘There is not any problem but it is very loose when we work hard and walk it’s will shrink and become loose when we tie it, the leg will be swollen and it will be tight again for that we will take it off if hurting or affect anything it’s just hard to work when it is loose. If there is an inflammation, it will be painful’ (Sarik, Client, Lines 69-72)*.

These issues appeared to be magnified when participants had to work harder or for longer hours, as described by Vuthy and Kheang:

> *‘It’s hot and itchy when I work a lot’ ‘No, I cannot wear it only when I cannot put it on. Sometimes, my leg was swollen and could not put it into (socket)…’ (Vuthy. Client, Lines 53 and 64-65)*.

> *‘Sometimes I just wish to return early so I can take the prosthetic leg off because it hurts’ (Kheang, Lines 132-133)*.

The issues described here are common to many prosthetic users around the world. Due to the heavy reliance on the limb for activities of daily living and income generation, being unable to wear the prosthesis could have a major impact on these participants’ lives. Access to the services that support clients to manage ongoing prosthetic use is explored in the next category of themes.

### Category 2: Access and use of prosthetic services

Due to their heavy reliance on the prosthesis and the need for it to fit and function well over time, participants described that access to prosthetic services was vital for enabling them to use their limbs. Three themes explore the issues surrounding access and use of services.

#### Theme 2.1: Difficulties attending and locating services

Many participants talked about difficulties of accessing prosthetic services and these experiences were often related to their initial contact with the clinic. These issues appeared to be related to i) a lack of awareness of the limb centre or service, ii) no clear understanding of what services were provided, and iii) not knowing where the centre was located. Participants described finding out about where to go to get their first prosthetic limb from other prosthetic users rather than from health care workers through a more traditional referral route following amputation surgery:

> *‘Someone who knows this centre told me when I lost my leg to come here and then if I have problems, that villager who works asked about here or there, telling me that the NGO will contact us’ (Sovy, Client, Lines 99-101)*.

Even for more established limb wearers, knowing where clinics were located was problematic when services changed or relocated. Participants described travelling to clinics that had closed or relying on local taxi drivers to direct them to their destinations. Ty describes how these problems of locating services had cost him more money for transport:

> ‘I had never come. Previously, I was told this organisation had finished, so then I went to XXXXX. Several days ago, I went to XXXXXX by motor taxi, which cost me about forty thousand Riels. When I reached there I was told my name was erased from their system due to I have my living address in XXX.”. (Ty, Client, Lines 192-198).

In addition to locating the centre, travelling there was described as a challenge for some clients. Being reliant on commonly used modes of transport in Cambodia, such as motorbikes, made travelling to centres difficult for some people, especially those with a transfemoral prosthesis, or when travelling longer distances:

> *‘I could not ride. I am afraid that the prosthetic leg could not bend to the desired position’ (Ty, Client, Lines 163-168)*.

Alongside the challenges in accessing services described above, many participants also talked about financial barriers to access. These will be described in the next theme.

#### Theme 2.2: Financial barriers to accessing prosthetic services

All participants in this study were recruited from the same INGO. Despite the service reimbursing travel and accommodation costs, participants reported that the cost of attending prosthetic services was a key barrier to access. Participants discussed how money was needed to travel to the centre, as well as pay for accommodation costs while away from home:

> ***‘****Because when I called, they made an appointment, but I did not have money to come, and I just delayed it until now, I just have money to come’ (Kheang, Client, Lines 78-80)*.

This financial impact appeared to be compounded by not being able to earn any income when attending the centre. This was thought by family members to be particularly challenging for people with a disability:

> *‘Because the majority of the people did not have money for transportation, and if so, it’s easier and saves them time and money on transportation’ (Sarum, Family, Lines 94 – 96)*.

Each of the access issues described above had a greater impact on participants who had to travel longer distances to the centre. These considerable barriers to accessing prosthetic services appeared to lead participants to adopt a crisis-management approach to maintaining limb use.

#### Theme 2.3: Crisis management approach to prosthetic use

Despite describing changes in the volume of residual limbs and the need for maintenance of prosthetic limbs to keep them in good working order, participants appeared only to attend the limb centre if their prosthesis was broken or too uncomfortable to wear:

> *‘If it was broken, I had to return’ (Thorn, Client, Line 84)*.

> *‘I must get their help when my leg had problems, I cannot live without my leg’ (Rany, Client, Line 128)*

To delay revisiting the centre, participants also described how they would repair the prosthesis themselves:

> *‘There is, sometimes the strap is broken, so I need to tie it with a metal wire’ (Kheang, Client, Line 161)*.

Other participants discussed managing socket fit and prolonging visits to the clinic through additional layers of socks:

> ‘*Just last time for socks and I told them it was loose, I was told to wear two layers of socks and wait for repair. Now, not three, I use four or five layers (Ratanak, Client, Lines144-145)*.

However, if there were no problems with the prosthesis or its fit, some participants would continue using it until it was completely worn out:

> *‘Never had problem, it fitted very well but now it is worn out with holes, broken and loose. When the clinicians looked at it, they said I need to have a new one (Souy, Client, Lines 77 – 78)*.

It appeared that this resilience and perseverance in terms of struggling on with a poor-fitting or broken limb until unable to function often led to a crisis point where participants could not undertake activities of daily living or carry out their paid employment:

> *‘If it is hard to walk only then I only come to the rehabilitation centre if it can be walk with I will not come. If as the appointment unless the foot is broken about 2 year or so I will come to receive the service and changing new foot is the most common because I walk a lot’ (Sarik, Client, Lines 82 – 84)*.

These times of ‘crisis’ seemed to be one of many examples of when prosthetic users needed to rely on other people for different types of support and experienced the broader impact of their disability on their standard of living and quality of life. These issues are illustrated in the final category of themes.

### Category 3: Sources of support and the impact on quality of life

Due to several of the issues raised in the themes above, it was found that participants in the study relied on a wide range of support mechanisms outside prosthetic services, such as immediate and extended family and the wider community. The following themes describe how the experiences of prosthetic use are embedded into the broader environment and have an impact on the individual’s entire social network.

#### Theme 3.1: Impact of disability on living conditions and reliance on financial support

Throughout the discussions, both clients and family members referred to ‘hard’ or ‘difficult’ living conditions and reported intermittent or continuous financial difficulties. In some cases, direct links were made between living with a disability, or the use of a prosthesis, and their standard of living:

> ***‘****Living condition is harder than before if I still have a leg, if I still have a leg I can go to work and have income for the home, but when I got amputated like that, sometimes I can go to work, but sometimes I can’t’ (Ratanak, Client, Lines 14-16)*.

Siha, a daughter of a client, discusses how she felt that a lot of people attending the centre for prosthesis rehabilitation would have financial difficulties:

> ***‘…****because financially it is not easy for everyone who comes here, not all of them have money because if there is disability, it will be hard to earn money. (Siha, Family, Lines 146-147)*.

Participants described how these difficult living conditions led them to be reliant on financial support from their families, such as money for food.

> ***‘****We are quite poor, even though I have a lot of children, but they need to support their family as well. They can’t always give us…I called to ask from them and when they give a little amount and I just use that to deal with the current condition that I am facing’ (Ing, wife of Kheang, lines 9-11)*

Among older participants, financial support was required to supplement military pensions. This is described by Eoun, who is a retired soldier:

> *‘In short, it is also difficult because the pension fund is only around 570000 riels per month, but I am also getting from the children, some of them don’t have to (Eoun, Client, Lines 17 - 18)*.

In most cases, it appeared to be children who provided this financial support, but some participants described having to rely on extended or different family members to send money, leading to further financial uncertainty:

> ‘… *just waiting for the relative sending some money to our parent for spending and buy some food’ (Kheang, Client, Lines 33-35)*.

Despite the necessity of this financial support, several participants emphasised the importance of working or of being independent and able to earn their own money. Vong, aged 50-59, describes a slight tension between wanting to work and his family wanting to support him as he is getting older:

> ***‘****Yes, they said I don’t need to do anything at this age and always comfort me, I want to go back to work if I am fully recovered. They always say no need, but I don’t want to feel too free, ….. Getting other people’s money to spend is not as easy as our own money. We feel sorry for the kids’ (Vong, Client, Lines 96-101)*.

#### Theme 3.2: Sources of social support from family and the community

In addition to financial provision, participants discussed a wide range of other forms of support received from their families or neighbours, which were often directly linked to the individual’s disability. For example, Sreysros describes the support she provides her husband:

> *‘… since my husband got this disability, I started to have trouble; everything needs to be done by myself. Because my husband has only one leg to rely on, thus I need to do everything by myself’ (Sreysros, Family, Lines 18-20)*.

It appears that receiving various types of social support from both family and the wider community was an essential part of being able to live with limb loss and cope when the prosthesis was not working well. For example, tangible forms of social support were in the format of the provision of transport (especially essential for getting to the clinic), help with activities of daily living, and helping with occupational activities, such as working on the farm. It was clear that this support was particularly essential when the participant was unable to use their prosthesis:

> ***‘****it’s just our families and neighbours when I don’t have prosthetic leg to use, they came to help, help to carry. They don’t discriminate at all they even help to encourage…(Sarik. Client, Lines 91-93)*.

Unsurprisingly, periods of illness or poor health were described as occasions when additional social support was required. This involved needing help to manage using their prosthesis or moving around their home:

> *‘There are helps from them when I did not feel well, I was helped to doff or don. When I feel well, I do it by myself.’ (Souy, Client, Lines 115-116)*.

Although participants described the closeness of their family unit and how they ‘managed’ or ‘solved’ problems for themselves, Siha explains how the changing support needs of her father can impact on the family who look after him:

> *‘but the importance thing is, he just quite old and he already disable, if it impacts on his health then it would also impact on the family who look after him, that’s it, just worry about his health’ (Shia, Family, lines 59-61)*.

The wider practicalities and impact on family life were highlighted by Sarik who describes the need for his family to give him lifts as modifications to the family mode of transport would make it unsuitable for other people to use:

> *‘My child carrying me I can ride it but it hard to take a brake if we modify the brake it hard for my child to ride it when they want to go around so they carrying me if bike-cycle I can ride it but it is difficult and tired because it far’ (Sarik, Client, Lines 62- 44)*.

## Discussion

This study explored the perspectives of people with lower limb amputation and their family members regarding access to prosthetic and rehabilitation services in Cambodia. Building on previous qualitative research in this context [5, 26], our findings provide deeper insight into how prosthetic services are accessed and used, and how these experiences are shaped by social, cultural, environmental, and economic factors. This reinforces the value of qualitative research in understanding service access within specific contexts and in informing the development of fit-for-purpose prosthetic devices and rehabilitation services in LMICs [8,9,14,18,20,21,24].

Across the findings, prosthetic devices and services were described as essential to daily life. Participants emphasised that prostheses enabled mobility, independence, employment, and the ability to fulfil family responsibilities, outcomes that extend well beyond physical function. These findings are consistent with previous studies in low-resource settings that describe prostheses as indispensable for survival and social participation [5,22,23].

Importantly, this reliance highlights the critical consequences of disrupted access to services, as periods without a functional prosthesis directly affected income, independence, and dignity.

Long intervals between visits characterised access to services, extended prosthesis use without repair or replacement, and a tendency to seek care only at points of crisis. While this pattern reflects the high value placed on prosthetic devices, it also exposes vulnerabilities within current service models. Participants described substantial barriers to accessing care, including travel distance, costs, and time away from income-generating activities, obstacles that have been widely reported in LMIC prosthetic service provision [11,14,36,37]. These access constraints appeared to contribute to risk-taking behaviours, such as prolonged use of damaged or uncomfortable prostheses, potentially increasing the risk of sudden device failure and loss of mobility.

Financial barriers continued to influence service use despite the availability of reimbursement mechanisms. These findings align with broader evidence showing that people with disabilities are disproportionately affected by poverty and that the costs associated with accessing rehabilitation can reinforce a ‘disability–poverty spiral’ [38,39]. Given the lifelong need for prosthetic review, repair, and replacement following amputation [37,40], patterns of infrequent service engagement observed in this study may have cumulative long-term consequences for health, participation, and economic security.

Participants’ experiences also highlighted the importance of prosthesis quality, durability, and suitability for the local environment. Reliable devices requiring minimal maintenance were highly valued, reflecting rugged terrain, physically demanding work, and limited access to routine repairs. Similar concerns have been raised in other LMIC settings, where prosthetic quality has been linked to limb abandonment and reduced service engagement [2,11,23,35]. These findings underline the need for service models and technologies that prioritise durability and environmental appropriateness alongside access.

Finally, this study demonstrates that experiences of accessing prosthetic services are embedded within wider family and community contexts. Family members played a crucial role in providing financial, practical, and emotional support, particularly during periods when participants were unable to use their prosthesis. While reliance on family support may be experienced as burdensome, it also enabled continued access to services and survival, echoing findings from other LMIC studies [23,43]. Including family perspectives, therefore, adds an essential dimension to understanding access to prosthetic and rehabilitation services and has implications for the design of services that are responsive to users’ broader social realities.

## Data Availability

All data produced in the present study are available upon reasonable request to the authors

## Acknowledgements

The authors are grateful to the funders, the Engineering and Physical Sciences Research Council (EPSRC) / National Institute for Health Research (NIHR) Global Challenges Research Fund (grant EP/R014213/1), and the Royal Academy of Engineering (RAEng grant RF/130).

The authors thank Exceed Worldwide, in particular Audrey Harte and Sam Simpson of the Exceed Research Network (ERN), for providing a critical review. We also thank the University of Southampton’s Institute for Life Sciences / FortisNet interdisciplinary musculoskeletal research network for supporting our preliminary work.

## Declaration of interest

We have no conflicts of interest relevant to this study.

## Notes

### Competing Interest Statement

The authors have declared no competing interest.

### Funding Statement

This study was funded by the Engineering and Physical Sciences Research Council, National Institute for Health Research Global Challenges Research Fund, grant EPR0142131, and the Royal Academy of Engineering, RAEng grant RF130.

### Author Declarations

Full ethical approval was granted by the Ministry of Health, National Ethics Committee for Health Research, Cambodia No. 311, NECHR and Faculty of Engineering & Physical Sciences at the University of Southampton ERGO45577.

